# Occupational Exposures to Blood and Body Fluids among Healthcare Workers in Ethiopia: A Systematic Review and Meta-Analysis

**DOI:** 10.1101/2020.05.20.20107953

**Authors:** Biniyam Sahiledengle, Yohannes Tekalegn, Demelash Woldeyohannes, Bruce John Edward Quisido

## Abstract

**Background:** Occupational exposure to blood and body fluids is a major risk factor for the transmission of blood-borne infections to healthcare workers. There are several primary studies in Ethiopia yet they might not be at the national level to quantify the extent of occupational blood and body fluid exposures among the healthcare workers. This systematic review and meta-analysis aimed to estimate the pooled prevalence of occupational blood and body fluid exposure of health-care workers in Ethiopia.

**Methods:** PubMed, Science Direct, Hinari, Google Scholar, and the Cochrane library were systematically searched; withal, the references of appended articles were also checked for further possible sources. The Cochrane Q test statistics and I^2^ tests were used to assess the heterogeneity of the included studies. A random-effects meta-analysis model was used to estimate the lifetime and 12-month prevalence of occupational exposure to blood and body fluids among health-care workers in Ethiopia.

**Results:** Of the 641 articles identified through the database search, 37 studies were included in the final analysis. The estimated pooled lifetime and 12-month prevalence on occupational exposure to blood and body fluids among healthcare workers were found to be at 54.15% (95% confidence interval (CI): 47.54–60.75) and 44.24% (95%CI: 36.98-51.51), respectively. The study identified a variation in healthcare workers whom were exposed to blood and body fluids across Ethiopian regions.

**Conclusion:** The finding of the present study revealed that there was a high level of annual and lifetime exposures to blood and body fluids among the healthcare workers of Ethiopia.

## Introduction

Occupational exposure to blood and body fluids (BBFs) is a major risk factor for the transmission of blood-borne infections to healthcare workers (HCWs). These exposures can heighten the risk of infection to Human Immunodeficiency Virus (HIV), Hepatitis B, and Hepatitis C. In many cases, exposures occur through a needle stick or contact in the eyes, nose, mouth, or when a broken skin comes in contact with the patient’s blood or body fluid [1,2]. According to the World Health Organization (WHO), it is estimated that about 3 million HCWs are exposed to bloodborne pathogens each year – occupational exposure causes approximately 170,000 to HIV infections, 2 million to HBV infections, and 0.9 million to HCV infections [3]. A recent review also stipulated that the prevalence of infections, such as HCV is significantly higher in HCWs than in the general population [1]. And more than 90% of HIV, HBV, and HCV exposures transpire in the developing countries [4].

Antecedent literatures publicized that blood or blood products accounted for 66% of exposures and the remaining involved vomits, urines, amniotic fluids, and cerebrospinal fluids; saliva; sputum; and other BBFs; 28% of these cases were fluids which contained visible bloods, and in some exposure incidents, multiple body areas were splashed or sprayed with these body fluids. The face is the most common exposure site reported: healthcare workers’ eyes (conjunctiva) were exposed to BBF (53%) of all reported cases. The mucosa of the mouth and nose were exposed in 11% and 5% of cases, respectively [5,6,7].

In sub-Saharan Africa, HCWs are at consequential risk of infection from blood-borne pathogens because of the excessive prevalence of such blood-borne infections in the general population [5,8]. A systematic review conducted in 21 African countries found a high prevalence of occupational exposures to blood and body fluids among HCWs – about two-thirds were exposed during their entire career, and almost half of them were exposed each year [9]. Additionally, evidence from every region of Africa indicates considerable variations in the prevalence of blood and body fluid exposures. The 12-month prevalence of all the types of occupational exposure to blood and body fluids ranged from 17.0% to 67.6% in Kenyan and Burundian studies. The estimated pooled 12-month prevalence was 48.0%. Regional pooled estimates covered from 33.9% to 60.7% in Southern Africa and Northern Africa [9].

In Ethiopia, occupational exposure to BBFs is a pressing concern and continues to have a significant problem in its healthcare system [10-14]. Antecedent studies also reported that standard precaution practices among HCWs were suboptimal, and the lack of compliance with these measures is still a great lookout [11, 13, 15, 16]. Though attention is paid to the safety of HCWs through the National Infection Prevention and Patient Safety (IPPS) initiatives, the number of exposures to BBFs reported did not manifest a sign of decline as evidenced by some studies [11,14,15,18].

Several primary studies in Ethiopia conveyed a high prevalence of occupational exposures to BBFs. However, the results were inconsistent [11,16-23]. For some instances, in Central Ethiopia, the prevalence of a 12-month BBF exposures among HCWs was 19.9% [11] and 41.3% [20]; in North Ethiopia 60.2% [16] and 31.7% [21]; and in East Ethiopia 43.8% [17] and 20.2% [14]. Currently in Ethiopia, no report exists to quantify the pooled prevalence of BBF exposures among its HCWs, even the existing review determined the prevalence of needle stick injury and did not estimate BBF exposures [24]. Moreover, the epidemiology of blood-borne infections in Ethiopia is on the rise and dynamically changing over the past decades, along with poor compliance toward standard precautions among HCWs. Given these developments, it is timely and crucial to investigate the burden of occupational BBF exposures among HCWs. Therefore, the objective of the present systematic review and meta-analysis directs to estimate the pooled prevalence of BBFs among HCWs in Ethiopia.

## Methods

This systematic review and meta-analysis were conducted subsequent to “the Preferred Reporting Items for Systematic Reviews and Meta-Analyses (PRISMA)” guidelines [25]. Studies were favoured according to the criteria outlined below (**Appendices A**).

## Eligibility Criteria

### Study Designs and

In this review, we appended cross-sectional studies and baseline assessment of longitudinal studies. Studies that reported the lifetime and/or 12-month prevalence of occupational exposure through blood and/or body fluid exposures to mucous membranes and broken skins were eligible to be included in the present review. Systematic reviews, letters to editors, short communications, qualitative studies, case series, case-control studies, and case reports were excluded. Also, articles that were not fully accessible, unsuccessful two-email contacts with the primary/corresponding authors were excluded, too. In addition to the aforementioned, studies restricted to HCWs’ needle stick and/or sharp injuries were excluded when data were not provided separately for blood and body fluid exposures. Lastly, the aggregate reports for blood and/or body fluid exposures and needle stick and/or sharps injuries were debarred from the study.

### Participants

Studies who met the following criteria were considered for inclusion:

Population: Healthcare works (HCWs) with direct contact to patients or blood/body fluids. We also encompassed studies which were conducted on a specific segment of the healthcare workforce (such as physicians, nurses, midwives, laboratory technicians, and cleaners).

Exposure: study examines occupational BBFs exposures

### Study Period

No restriction on publication date.

### Language

Articles which were only reported in the English language.

### Article Searching Strategy

MEDLINE/PubMed, Hinari, Science Direct, and the Cochrane Library databases from inception until January 31, 2020, that reported the prevalence of occupational exposures to blood and/or body fluids among HCWs in Ethiopia were sought. Literature search strategies were developed using medical subject headings (MeSH) and text words related to occupational blood and/or body fluid exposures. The following search terms were used and combined using Boolean operators: “prevalence”, “magnitude”, “occupation”, “exposure”, “accident”, “occupational exposure”, “accidental exposure”, “accidental occupational exposure”, “occupational disease”, “occupational hazard”, “cross-infection”, “blood”, “body fluid”, “blood spill”, “blood-borne pathogens”, “blood-borne infection”, “health-care workers”, “health workers”, “medical personnel”, “health personnel” and “Ethiopia”. The electronic database search was also supplemented by searching for gray literature through Google scholar, Google searching, and Ethiopian University digital repositories (such as the Addis Ababa University Digital Library). To ensure literature saturation, the reference list of appended studies and/or relevant studies identified through the search were scanned as well. Finally, the literature search was limited to the English language and human subjects (**Appendices B)**.

## Operational Definition

### Occupational Blood and Body Fluid Exposure

In this review, “occupational blood and body fluid exposure” is defined as any exposure to potentially harmful biological agents or infectious materials that occurs as a result of one’s occupation, which includes the patient’s blood and other body fluids. We appended studies that reported the lifetime or 12-month prevalence of occupational exposure through blood and body fluid contacts from at least one of these routes (eye, mouth, mucous membrane, and non-intact skin).

#### Healthcare Workers

Healthcare workers (HCWs) are referred to as paid or unpaid individuals (eg: full-time employees or medical students) working in a health-care setting whose activities involve direct contact with patients, or with blood or other body fluids from the patients. Hence, we incorporated studies which involved physicians, nurses, midwives, health officers, laboratory technicians, anesthetists, auxiliary healthcare workers, residents, or interns undertaking clinical training or gaining experiences in the health-care settings.

### Study Selection and Data Extraction

In this review, all the searched articles were imported into the EndNote version X^4^ software, and after that, the duplicate articles were removed. Two investigators (BS and YT) independently screened and identified articles by their titles, abstracts, and full-texts to determine eligibilities against predetermined inclusion and exclusion criteria. Afterwards, the screened articles were compiled together by the two investigators, and discrepancies were resolved through unanimous consensus.

The data extraction form was prepared using Microsoft Excel Spreadsheet. Two reviewers extracted data from the studies and were entered into Microsoft Excel. The data extraction form included: (i) name of primary author; (ii) year of publication; (iii) region; (iv) sample size; (v) study population; (vi) type of study design; (vii) sampling technique (viii) response rate and (ix); 12 months and lifetime prevalence of blood and body fluid exposure among HCWs.

### Quality Assessment

The qualities of the appended studies were assessed and the risks for biases were judged using the Joanna Briggs Institute (JBI) quality assessment tool for the prevalence studies [26]. There were nine parameters: (1) appropriate sampling frame, (2) proper sampling technique, (3) adequate sample size, (4) study subject and setting description, (5) sufficient data analysis, (6) use of valid methods for the identified conditions, (7) valid measurement for all participants, (8) using appropriate statistical analysis, and (9) adequate response rate (adequate if 60% or higher). Failure to satisfy each parameter was scored as 1 if not 0. The risks for biases were classified as either low (total score: 0 to 2), moderate (total score: 3 or 4), or high (total score: 5 to 9). Two reviewers (BS and YT) assessed the quality of the studies included. Finally, articles with scores of 5 to 9, which meant having a high risk of biases were debarred (**Appendices C**).

## Statistical Analysis

Primarily, appended studies were categorized whether they have measured the lifetime prevalence of blood and body fluid exposures or whether they are on a 12-month prevalence, and later were entered into the STATA version 14. The meta pop program was utilized to estimate the pooled prevalence of lifetime and 12-month prevalence of blood and body fluid exposure among HCWs. Accordingly, the prevalence of blood and body fluid exposure (p) were estimated using data from the appended studies which reported the proportion of HCWs whom were exposed to body fluids at any time during their career, and twelve-month prevalence was appraised using data from the studies which reported the proportion of participants exposed to body fluids in the preceding 12 months. Corresponding standard errors (SE) were calculated using se = √p(1 - p)/n. The researchers estimated the pooled prevalence of blood and body fluid exposures using random-effects meta-analysis based on the DerSimonian and Laird approach. The existence of heterogeneity among the studies was checked using the I^2^ test statistics. Heterogeneity will be classified into the following three categories: low heterogeneity (I^2^ index < 25%), average heterogeneity (I^2^ index= 25–75%), and high heterogeneity (I^2^ index > 75%). Also, a p-value of < 0.05 is used to declare heterogeneity. Thus, a random-effects model was used to analyze data in this study, since the estimated both 12 months and lifetime prevalence of BBFs was found to be high. Finally, Meta-regression analysis was used to evaluate the association between the prevalence of BBFs and publication year, and sample size in the selected studies.

## Publication Bias

In this meta-analysis, possible publication biases were visualized thru funnel plots. Symmetrical large inverted funnels resembled the absence of publication biases. Also, the probability of publication biases were tested using two main statistical methods (Egger’s and Begg’s tests) which were wield to test funnel plot asymmetries. The level of significance for asymmetries were viewed as p<0.05.

## Sensitivity Analysis

Also, sensitivity analyses were undertaken – the stability of the pooled estimate for each study. The investigation was done by excluding a single individual study from the analysis at a time to explore the robustness of the findings.

## Results

### Description of the Studies

The initial electronic searches generated 641 studies using international databases and Ethiopian university research repositories. The database included PubMed (82), Science Direct (61), Hinari (279), Google Scholar (196), Cochrane Library (1), and the remaining 22 studies were identified through manual search. Of these, 151 duplicates were identified and effaced. From the tarry of 490 articles, based on the pre-defined eligibility criteria, 428 articles were excluded after reading their titles and abstracts. 62 full-text articles remained and were further assessed for their eligibilities. Finally, based on the pre-defined inclusion and exclusion criteria and quality assessment, only 37 articles were extracted for the final analyses [10-14, 16-23, 27-50] (**Fig. 1**).

**Fig.1:**
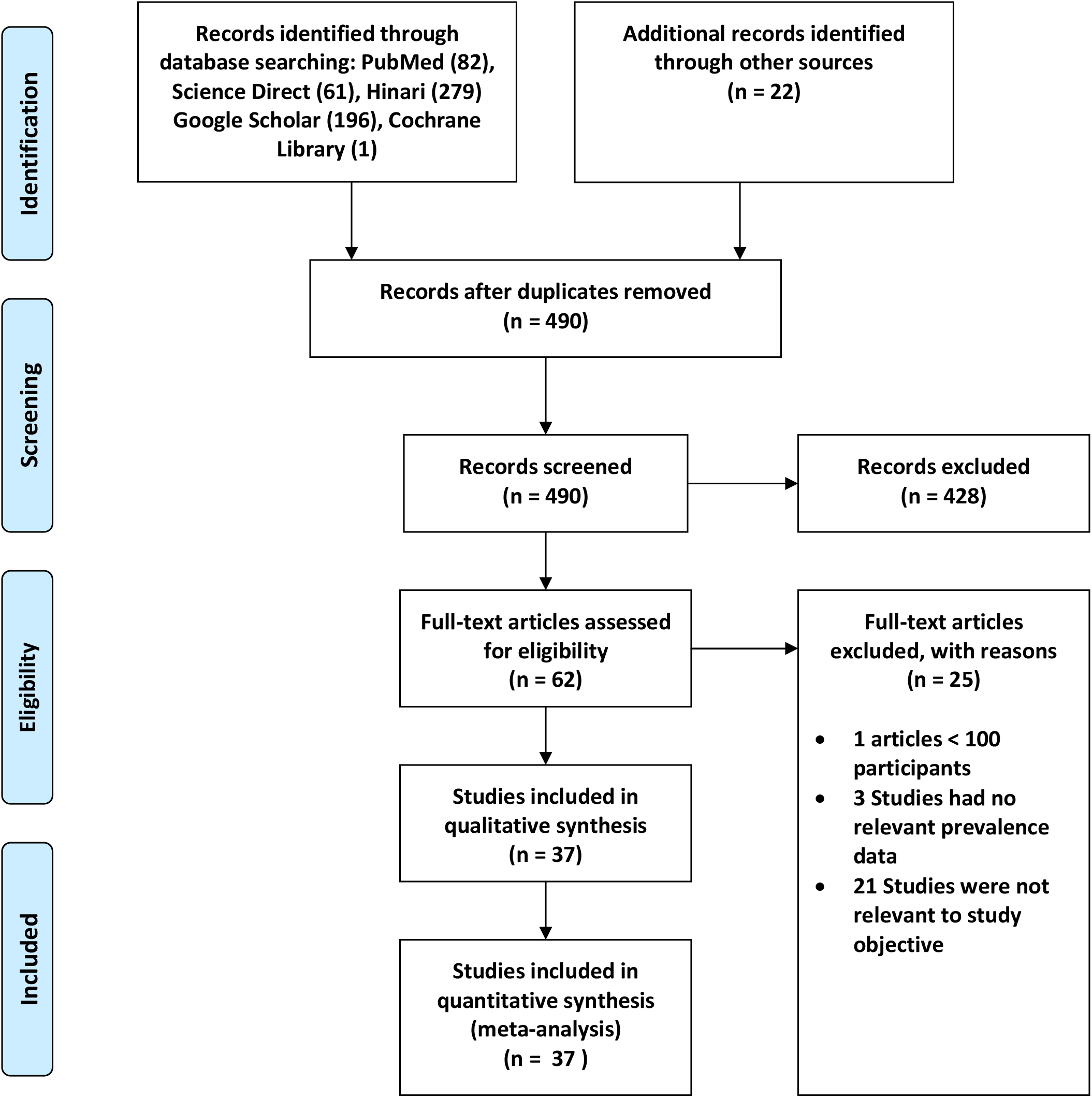
Flow diagram, systematic review, blood and body fluid exposure among healthcare workers in Ethiopia, 2007-2020

### Characteristics of the Appended Studies

The general characteristics of the favored articles were presented in **Table 1**. Of the 37 articles included in this review and meta-analyses, 14 were conducted in Addis Ababa; 10 in the Amhara Region; 6 in Oromia Region; 4 from the Southern Nations, Nationalities, and People (SNNP); 2 in Harari Region; and only 1 from Tigray Region. A total number of 11,168 healthcare workers participated in the study – the highest and lowest sample sizes were from the studies of Geberemariyam et al. [13] in the Oromia Region (648 HCWs), and [47] in Addis Ababa (104 HCWs). All the appended studies were cross-sectional studies. Twenty-three studies were conducted solely among hospital healthcare workers. Among the studies, twenty-three of them also presented data regarding 12-month prevalence on occupational exposures to BBFs [10-12, 14, 16, 17, 19-23, 27-29, 35, 37-39, 44-47, 49], and the lifetime prevalence on BBF exposures were reported in twenty-six studies [11-15, 18-20, 22, 28-34, 36, 39-43, 45, 46, 48, 50]. From the studies, thirteen articles have reported having both the 12-month and lifetime BBFs exposure prevalence [10-12, 14, 19, 20, 22, 28, 29, 39, 44-46]. The latest article was published in 2020 [10], and the earliest study was concluded last 2007 [45]. The prevalence of 12 months BBFs among the Ethiopian HCWs ranged from 16.5% [12] to 67.5% [23] in Addis Ababa Region. The lifetime prevalence of BBFs varied from 28.8% in the Harari Region [14] to 81.0% in the Amhara Region [33]. In this review, a low risk of bias was realized in 33 (89.2%) of the included studies (**Appendices C**).

**Table 1:**
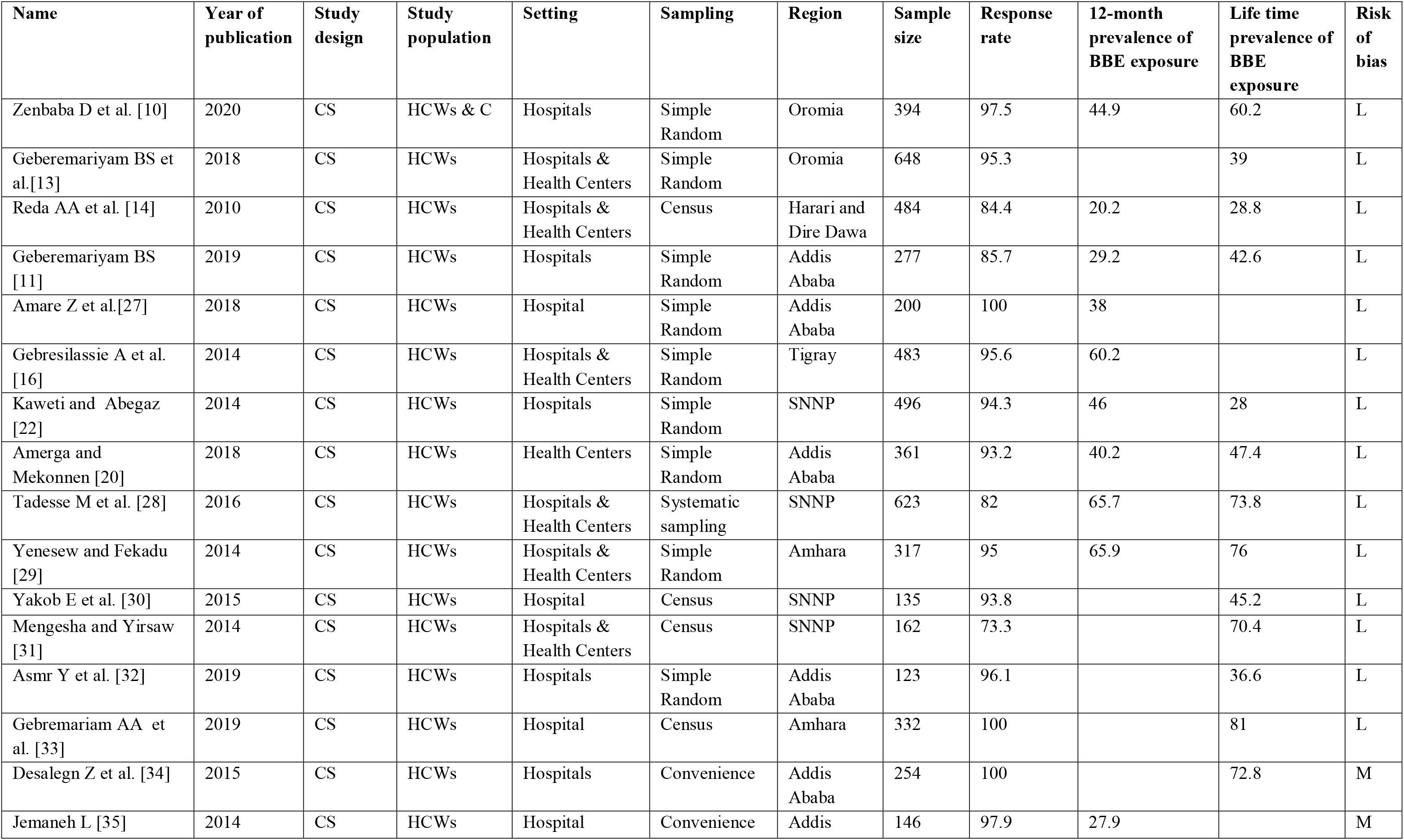
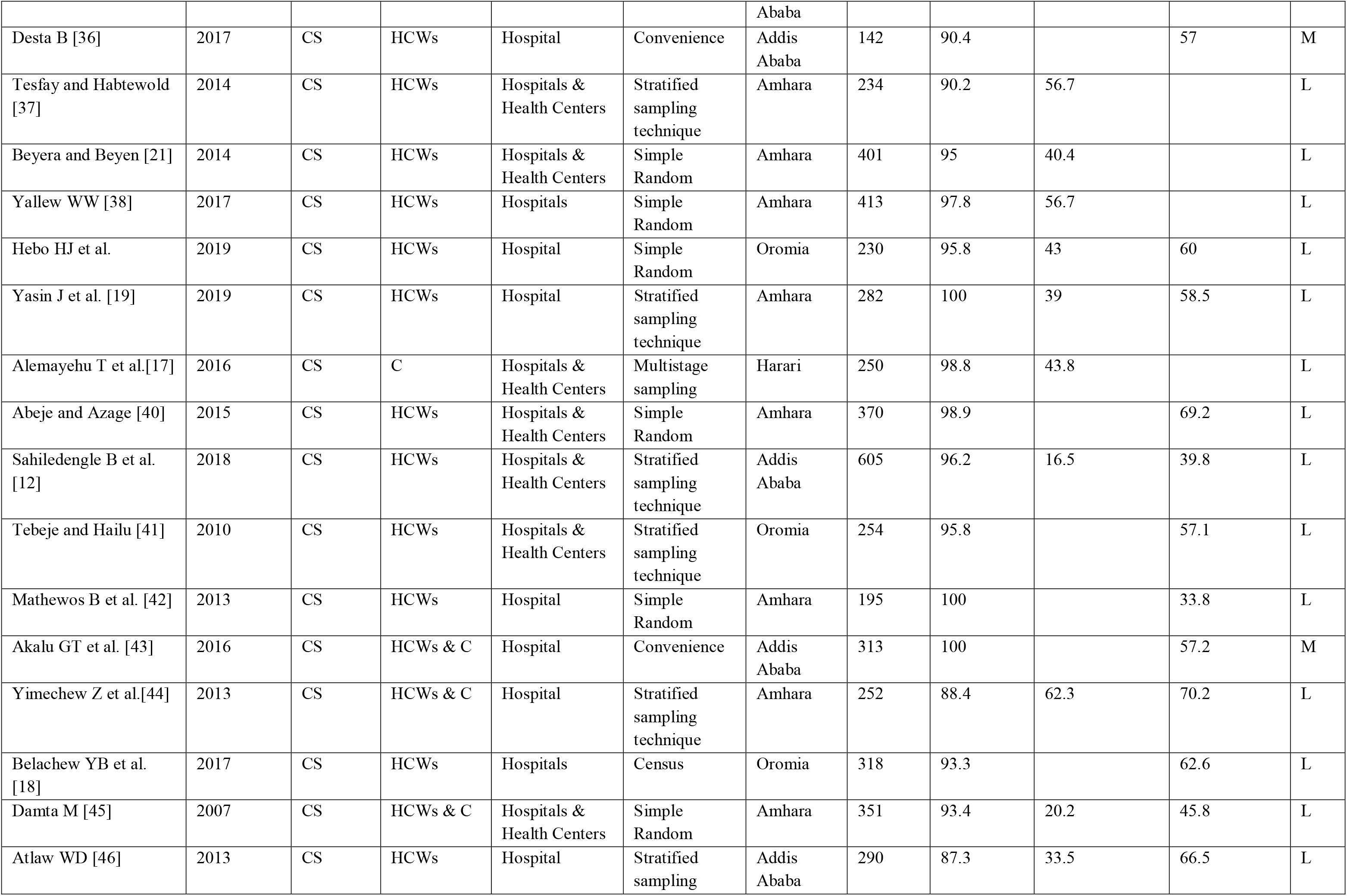
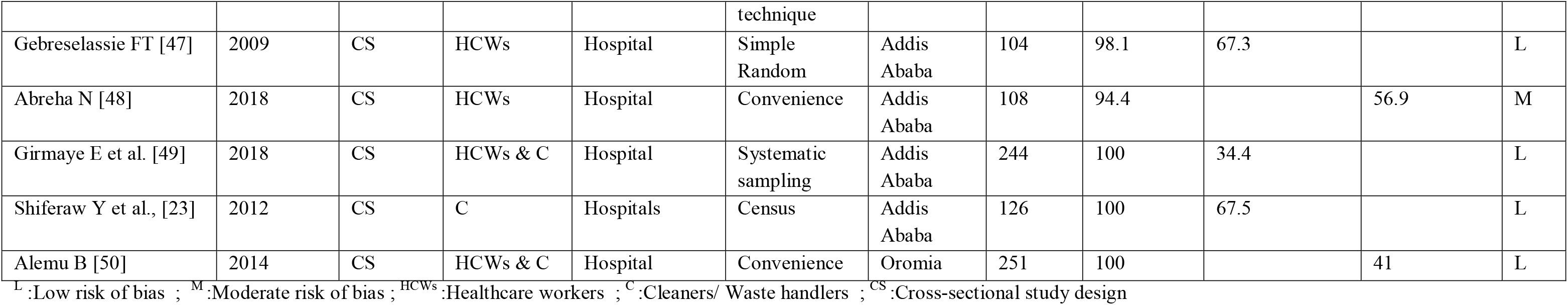
Studies identified in the systematic review on blood and body fluid exposure among health-care workers in Ethiopia, 2007–2020

### Prevalence of Blood and Body Fluid Exposures among HCWs in Ethiopia

The current meta-analysis using the random-effects model conveyed that the estimated overall pooled prevalence of 12 months BBF exposures among HCWs in Ethiopia was 44.24% (95%CI: 36.98-51.51) with a significant level of heterogeneity (I^2^ = 97.9%; p < 0.001) (**Fig. 2**). The lifetime pooled prevalence of BBFs using the random-effects model was 54.15% (95% CI 47.54-60.75) with a significant level of heterogeneity (I^2^ = 97.6%; p < 0.001) (**Fig. 3**).

**Fig.2:**
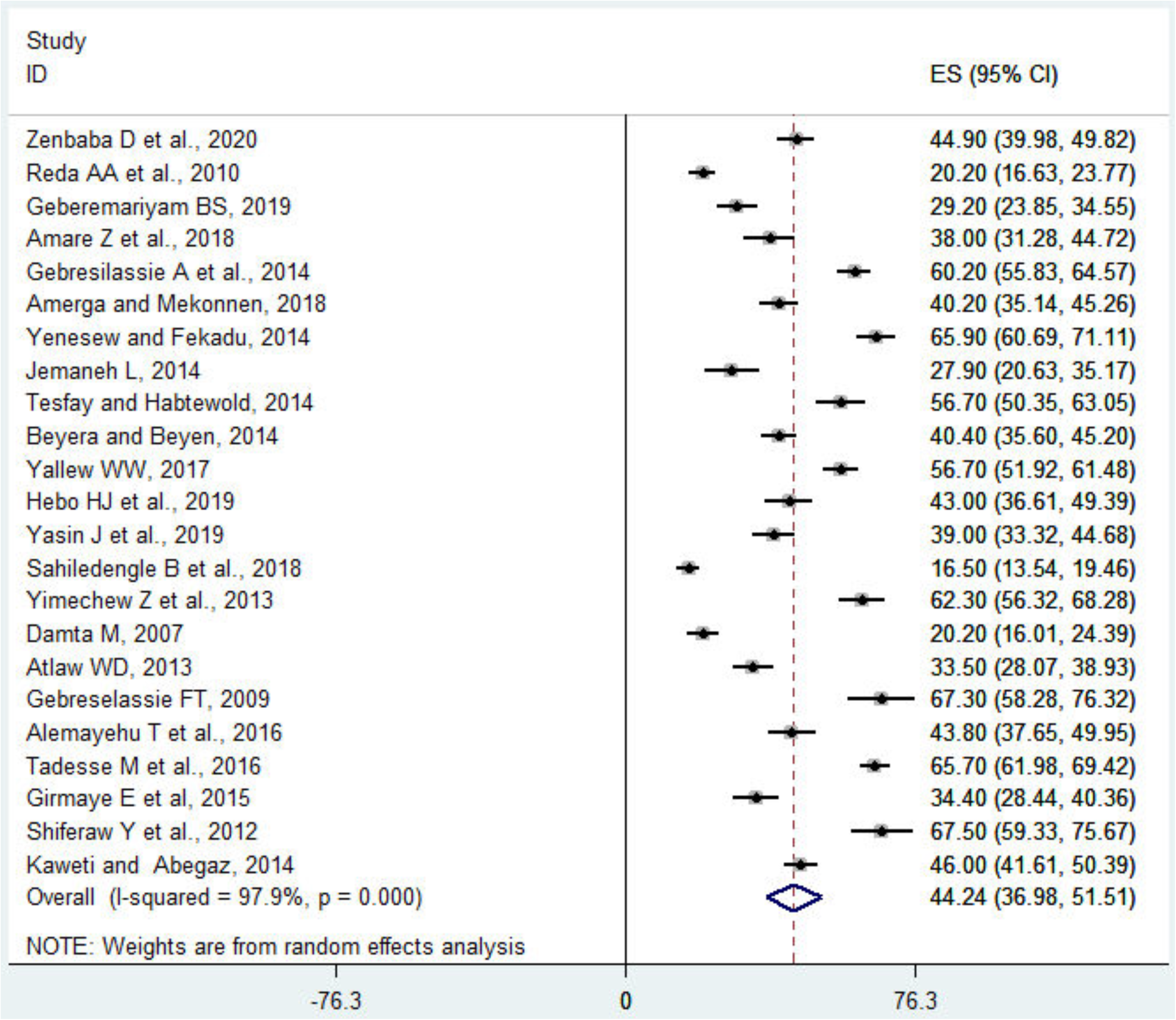
Meta-analysis, 12 month prevalence of blood and body fluid exposure among health-care workers in Ethiopia, 2007-2020

**Fig.3:**
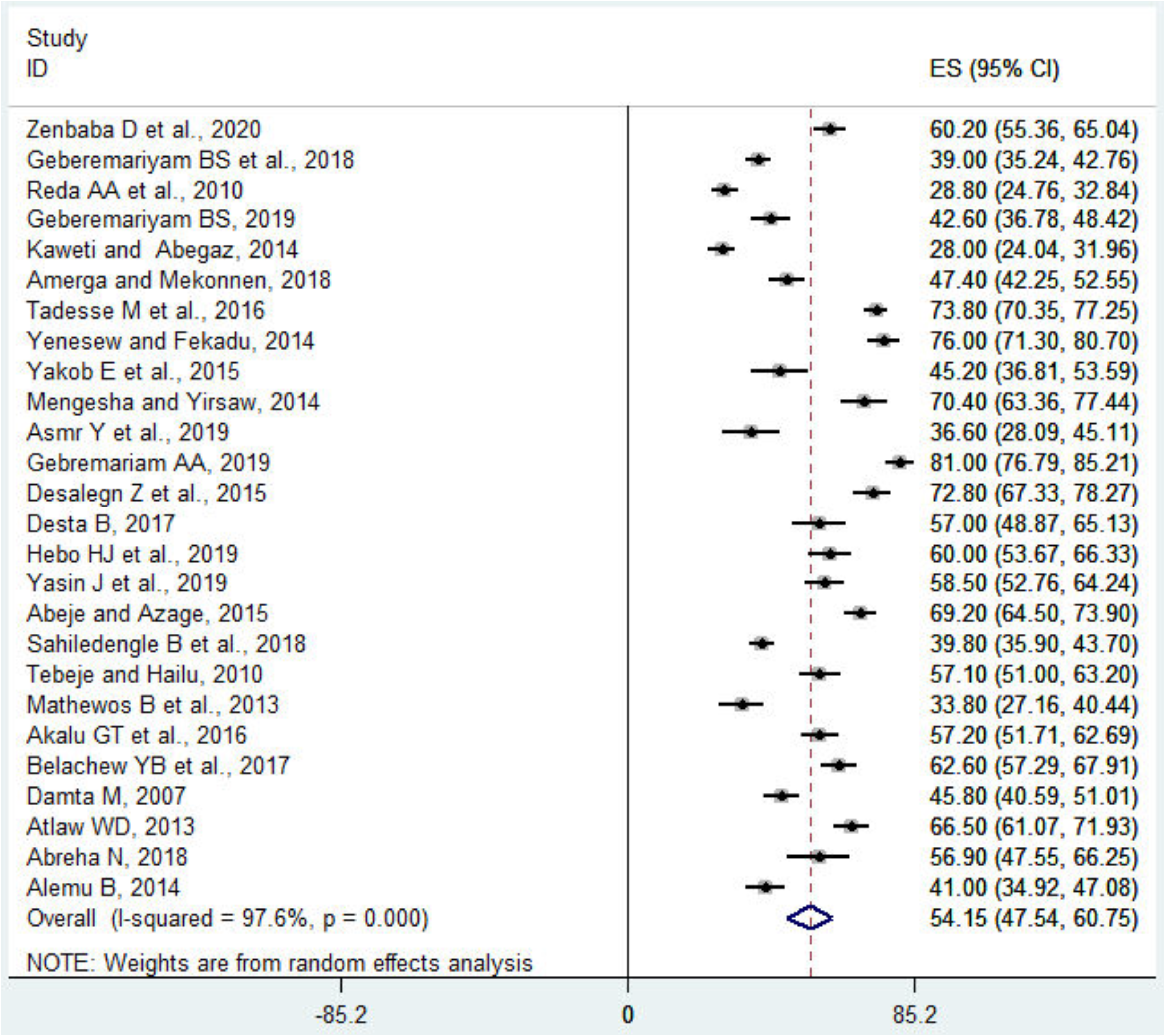
Meta-analysis, lifetime prevalence of blood and body fluid exposure among health-care workers in Ethiopia, 2007-2020

### Investigation of Heterogeneity and Subgroup Analysis

The included studies in this meta-analysis exhibited a statistically significant heterogeneity between studies (I^2^ = 97.9%; p < 0.001, and I^2^ = 97.6%; p < 0.001) for the 12-month and lifetime BBF exposure prevalence estimates, respectively. Accordingly, the random-effects model was used to adjust the observed variability. In identifying the possible source of heterogeneity, subgroup analyses were utilized based on the geographical regions, type of healthcare facilities, year of publication, and sample size. However, the level of heterogeneity between studies remained high after subgroup analysis (**Table 2**).

**Table 2:** Subgroup meta-analysis, blood and body fluid exposure among health-care workers in Ethiopia, 2007–2020

| Prevalence type | Variables category | Subgroup | Number of studies included | Sample size | Prevalence (95% CI) | Heterogeneity across the studies |  |
| --- | --- | --- | --- | --- | --- | --- | --- |
|  |  |  |  |  |  | I<sup>2</sup> (%) | P-value |
| Lifetime prevalence | Region | Addis Ababa | 9 | 2,473 | 53.00(44.47-61.53) | 94.7 | <0.001 |
|  |  | Oromia | 6 | 2,095 | 53.25(44.02-62.49) | 94.6 | <0.001 |
|  |  | Amhara | 6 | 1,847 | 60.83(47.03-74.62) | 97.7 | <0.001 |
|  |  | SNNP | 4 | 1,416 | 54.35(28.38-80.31) | 99.1 | <0.001 |
|  |  | Harari | 1 | 484 | 28.80 (24.76-32.83) | - | - |
|  | Type of healthcare facility | Hospital | 16 | 4,140 | 53.81(45.45-62.16) | 97.0 | <0.001 |
|  |  | Hospital & health centers | 9 | 3,814 | 55.50(43.15-67.84) | 98.5 | <0.001 |
|  |  | Health center | 1 | 361 | 47.40(42.25-52.55) | - | - |
|  | Publication year | 2007-2014 | 9 | 2,800 | 49.67(36.85-62.49) | 98.2 | <0.001 |
|  |  | 2015-2020 | 17 | 5,515 | 56.56(49.44-63.68) | 96.8 | <0.001 |
|  | Sample size | >=300 | 13 | 5,612 | 54.51(44.13-64.89) | 98.6 | <0.001 |
|  |  | <300 | 13 | 2,703 | 53.81(46.70-60.92) | 93.4 | <0.001 |
|  | Sampling technique | Probability | 21 | 7,247 | 53.47(45.85-61.10) | 98.0 | <0.001 |
|  |  | Non-probability | 5 | 1,068 | 57.02(45.74-68.29) | 93.1 | <0.001 |
|  | Risk of bias | Low | 22 | 7,498 | 52.91 (45.53-60.30) | 97.9 | < 0.001 |
|  |  | Moderate | 4 | 817 | 61.27(52.38-70.16) | 85.2 | < 0.001 |
| A 12-month prevalence | Region | Addis Ababa | 9 | 2,353 | 39.09(28.66-49.52) | 96.8 | <0.001 |
|  |  | Oromia | 2 | 624 | 44.19(40.29-48.09) | 0.0 | 0.644 |
|  |  | Amhara | 7 | 2,250 | 48.69(35.53-61.85) | 97.8 | <0.001 |
|  |  | Tigray | 1 | 483 | 60.20 (55.83-64.57) | - | - |
|  |  | SNNP | 2 | 1,119 | 55.89(36.58-75.19) | 97.8 | <0.001 |
|  |  | Harari | 2 | 734 | 31.86(8.73-54.98) | - | - |
|  | Type of healthcare facility | Hospital | 13 | 3,454 | 45.19 (38.55-51.83) | 94.5 | <0.001 |
|  |  | Hospital & health centers | 9 | 3,748 | 43.24(28.72-57.77) | 99.0 | <0.001 |
|  |  | Health center | 1 | 361 | 40.20(35.14-45.26) | - | - |
|  | Publication year | 2007-2014 | 12 | 3,684 | 47.21(36.50-57.92) | 98.0 | <0.001 |
|  |  | 2015-2020 | 11 | 3,879 | 41.04 (30.63-51.45) | 98.0 | <0.001 |
|  | Sample size | >=300 | 11 | 4,722 | 43.32(31.52-55.12) | 98.8 | <0.001 |
|  |  | <300 | 12 | 2,635 | 45.03(37.47-52.59) | 94.1 | <0.001 |
|  | Sampling technique | Probability | 22 | 7,417 | 44.97 (37.50-52.44) | 98.0 | <0.001 |
|  |  | Non-probability | 1 | 146 | 27.90 (20.63-35.17) | - | - |
|  | Risk of bias | Low | 22 | 7,417 | 44.97 (37.50-52.44) | 98.0 | <0.001 |
|  |  | Moderate | 1 | 146 | 27.90 (20.63-35.17) | - | - |

SNNP= South Nation Nationalities and Peoples

The prevalence of 12 months BBFs was found to be higher in the Tigray Region 60.20 % (95%CI: 55.83-64.57) and the least was reported from the Harari Region 31.86% (95%CI: 8.73-54.98). This meta-analysis also found that the lifetime prevalence of BBF exposures differed between various regions, and the highest prevalence was found in the Amhara Region, 60.83% (95%CI: 47.03-74.62), followed by SNNP Region, 54.35% (95%CI: 28.38-80.31), and finally, the least in Harari Region, 20.80% (95%CI: 24.76-32.83). Withal, the 12 months and lifetime prevalence of BBF exposures were 41.04 (95%CI: 30.63-51.45) and 56.56% (95%CI: 49.44-63.68) in studies published between 2015 and 2020, respectively (**Table 2**).

### Sensitivity Analysis

To identify the source of heterogeneity and to explore the robustness of the findings, a leave-one-out sensitivity analysis was employed. The result of sensitivity analyses using the random-effects model revealed that no single study influenced the overall prevalence of 12 months and lifetime BBF exposures among HCWs (**Appendices D**).

### The Publication Bias

The presence of publication bias was evaluated using funnel plots and Egger’s tests at a significance level of less than 0.05. The findings revealed that publication bias was not significant for the studies reported in the 12-month prevalence of BBF exposures (p = 0.05) (**Fig. 4**). In the same manner, it was not statistically significant (p = 0.92) for the lifetime BBFs exposures, as well (**Fig. 5**).

**Fig.4:**
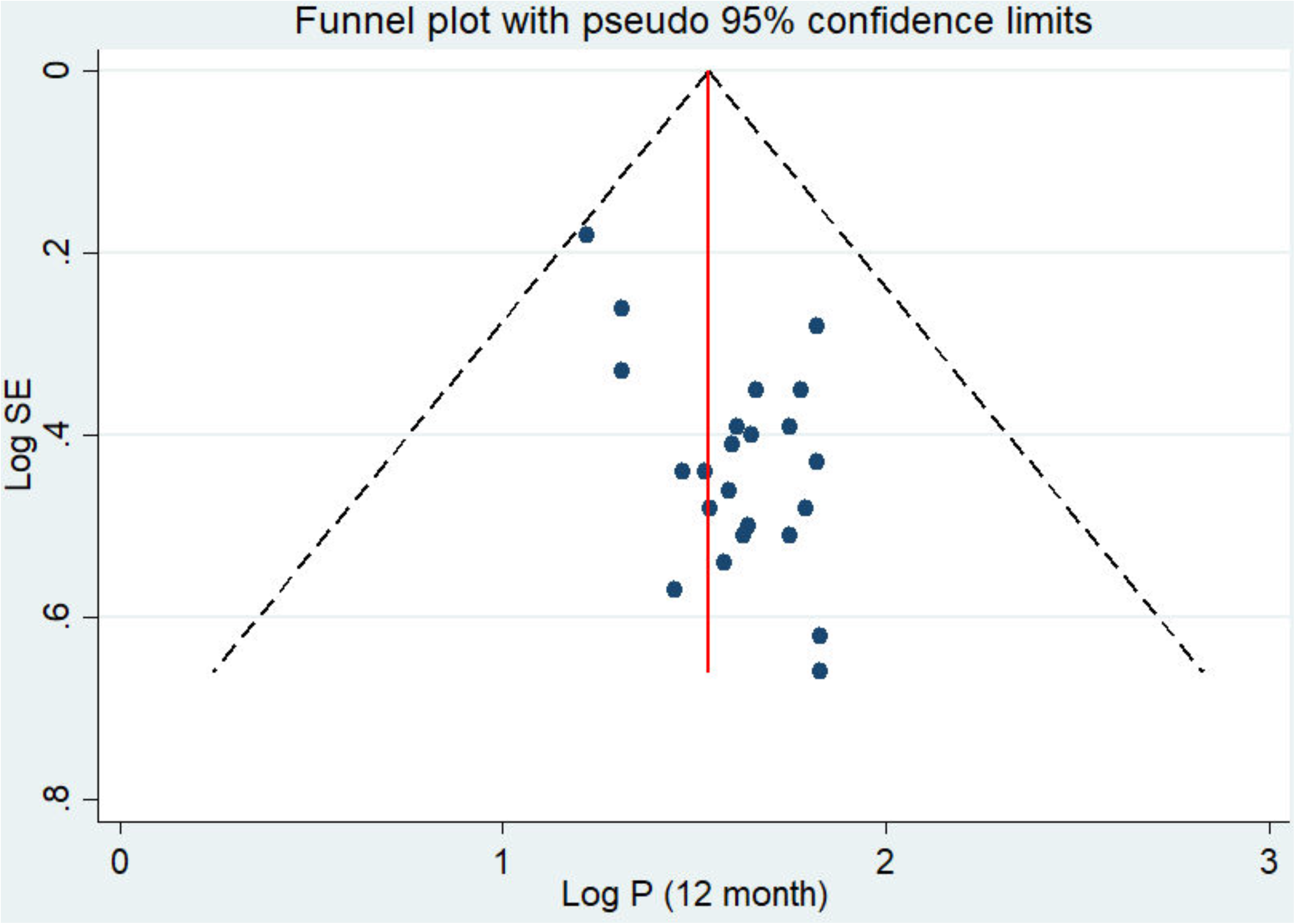
Publication bias of 12 month prevalence of BBFs exposure among HCWs in Ethiopia, 2007-2020.

**Fig. 5:**
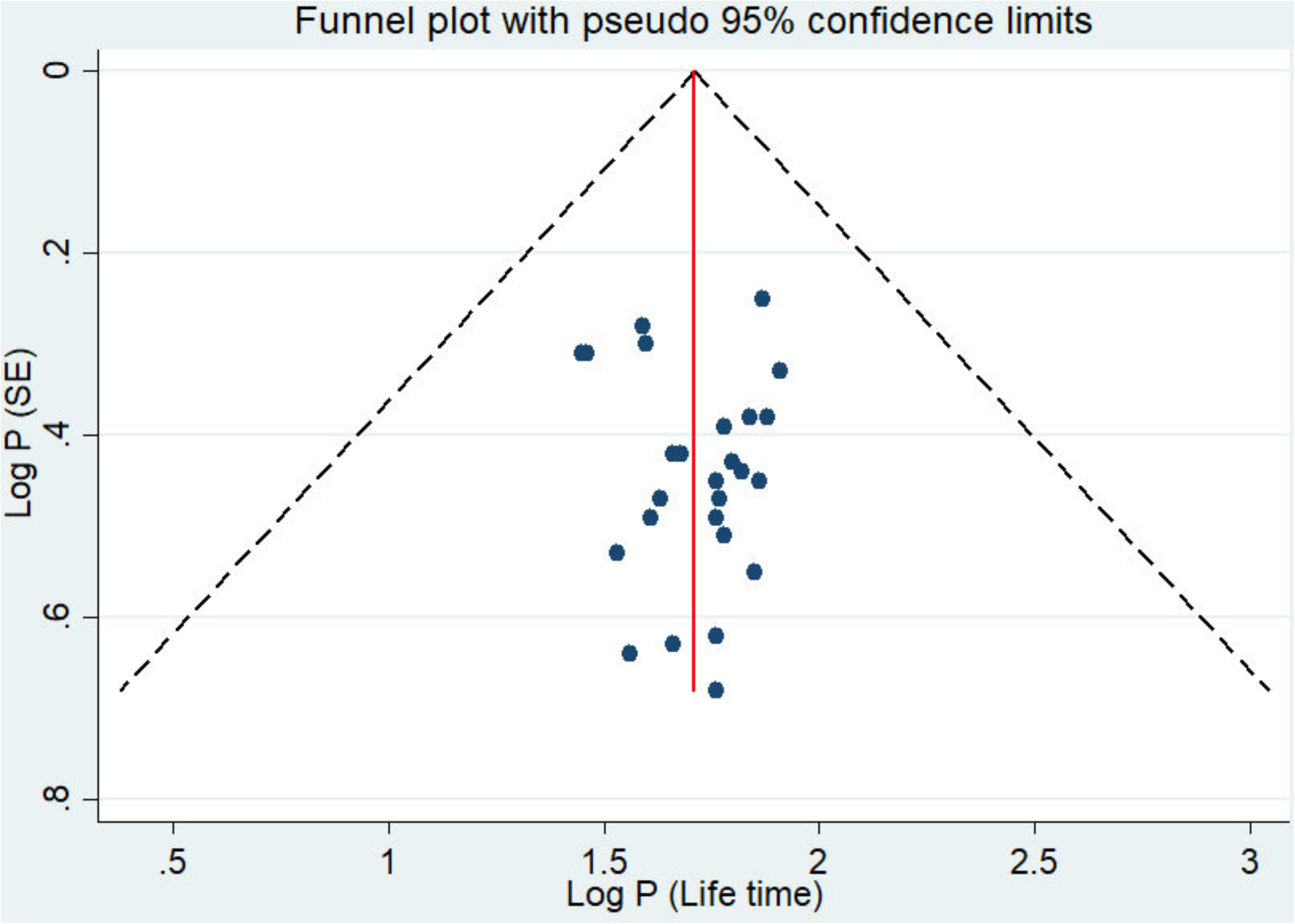
Publication bias of lifetime prevalence of BBFs exposure among HCWs in Ethiopia, 2007-2020.

### Meta-Regression Analysis

The results of the meta-regression analysis showed that the publication year and the sample size were not significant sources of heterogeneity. In this study, no significant relationship were identified between the 12 month prevalence of BBFs and publication year (p-value=0.76), and sample size (p-value= 0.44). Similarly, there was no significant association between the lifetime prevalence of BBFs and publication year (p-value=0.42) and sample size (p-value= 0.48) (**Table 3**).

**Table 3:** A meta-regression analysis of factors for heterogeneity of the prevalence of blood and body fluid exposure among the healthcare workers in Ethiopia, 2007-2020.

| Prevalence estimate | Heterogeneity source | Coefficients | Std. error | p-value |
| --- | --- | --- | --- | --- |
| 12 months | Publication year | -0.3185998 | 1.026952 | 0.76 |
|  | Sample size | -0.0190527 | 0.0242865 | 0.44 |
| Lifetime | Publication year | 0.7797819 | 0.940355 | 0.42 |
|  | Sample size | -0.0147593 | 0.020484 | 0.48 |

## Discussion

Each year, hundreds of thousands of HCWs, including waste handlers, face the risk of blood-borne diseases due to occupational BBF exposures [3, 8, 9, 51]. In Ethiopia, despite the recognition on the importance of HBV, HCV, HIV, and other diseases transmitted through BBFs by the Federal Ministry of Health (FMoH), currently, there is dearth of systematic reviews and meta-analyses that estimated the prevalence of BBFs exposure among HCWs. In this reckon, this study was the first systematic review and meta-analysis that aimed to estimate lifetime and a 12-month prevalence on occupational exposure to BBFs among Ethiopian HCWs. This review involved the results of 37 articles which investigated the prevalence of BBF exposures, and a high burden on occupational exposures to BBFs among HCWs in Ethiopia was evidently identified.

The estimated pooled 12-month and lifetime prevalence on BBF exposures among HCWs in Ethiopia were 44.2% and 54.2%, respectively. Forbye, the 12-month BBFs prevalence in the primary studies ranged from 16.5% [12] to 67.5% [23]. In parallel, the lifetime prevalence ranged from 28.8% [14] and 81.0% [33]. This 12-month pooled prevalence estimate was almost comparable from the pooled estimate from East Africa (47.3%) [9], Côte d’Ivoire, Mali and Senegal (45.7%) [52], and a study by Bi P et from Australia, revealed that 42% of HCWs had body fluid exposures in a year on their study [53]. However, it was lower than the studies conducted in Turkey (57%) [54] and Nigeria 67.5% [55]. These differences might be subjected to the variances in the socio-demographic, cultural characteristics of study participants, and study health facility setup variations.

This study explicated a higher prevalence of lifetime BBF exposures (54.2%), however, it was subservient than the reviews from the 65 studies in 21 African countries (65.7%) [9]. The foremost reason for this variation may be due to study setting dissimilarities. This finding is also inconsistent with a study in Iran which reported the prevalence of exposures at 46.47% [56]. The variance could be due to the discrepancies in the study participants, the type of healthcare facilities, and socio-demographic factors.

In this review, the researchers identified a variation in the HCWs’ exposure to BBFs across the Ethiopian regions. The lifetime (60.83% in Amhara Region) and 12 months (60.20% in Tigray Region and 48.69% in Amhara Region) occupational exposure to BBFs were consistently more frequent in Northern Ethiopia, and less in Harari Region (lifetime prevalence of 28.80% and 12-month prevalence of 31.86%). The probable rationale for these regional variations may be due to: the number of studies included; type of healthcare facilities; and geographical and demographical differences. The other possible vindication for these disparities may be partially explained by the polarities in the levels of standard precaution practices among the HCWs in the various regions. As one study reported, 80.8% of the HCWs regularly follow standard precautions in Eastern Ethiopia, including the Harari Region [14].

A laudative prevalence of BBF exposures among HCWs working exclusively in hospitals than those in the health centers (primary healthcare units) was also found. Almost half of the HCWs working in hospitals of Ethiopia had at least one BBFs exposure in their lifetime and in the last 12 months. The finding was predictable because these HCWs had higher workloads and they performed further medical procedures, which may have exposed them to occupational BBFs compared to those in the health centers. Therewithal, the high prevalence of BBF exposures among HCWs working in the hospitals had significant implications because most of the blood-borne viruses, such as HCV, HBV, and HIV, may haply spread through BBFs exposures, therefore, enhancing HCWs’ compliance towards standard precautionary measures is deemed necessary.

## Limitations

This review article had a few adversities due to its limitations. One of which was the cross-sectional design nature of the included studies and all were based on self-reported data whilst estimating the prevalence of occupational BBFs exposures. Additionally, social desirability and recall biases were likely present. Since the study was conducted in Ethiopia, included healthcare facilities and the generalization of the study findings was limited to these similar contexts. Further, there was no study obtained from the some Ethiopian regions, such as Afar Regional State and Benshangul-Gumuz Regional State and this might probably affect the generalizability of the present findings at a national level.

## Conclusions

This review exhibited a higher percentage of occupational exposures to BBFs among HCWs in Ethiopia. The available evidences suggest that more than two-in-five and one-half of healthcare workers in Ethiopia were exposed to BBFs annually and in their lifetime, respectively.

Therefore, efforts should be implemented to reduce the high burden of occupational blood and body fluid exposures through effective implementation of standard precaution measures along with aggressive occupational health and safety activities.

## Data Availability

All relevant data are within the manuscript and its supporting information files

**Abbreviations**
AOR: Adjusted odds ratio;
BBFs: Blood and body fluids;
HCWs: Healthcare workers;
CI: Confidence interval;
IPPS: Infection Prevention and Patient Safety;
PRISMA: Preferred Reporting Items for Systematic Reviews and Meta-Analyses;
WHO: World Health Organization

## Declarations

### Ethics approval and consent to participate

Not Applicable

### Consent for publication

Not Applicable

### Availability of supporting data

All relevant data are within the manuscript and its supporting information files.

## Authors’ Contribution

BS: Conceptualizes, design the study and data curation, performed the analysis, wrote and approved the final manuscript. YT: Data curation and performed the analysis, and approved the final manuscript DW: Contribute to the analysis, critically reviewed the manuscript and approved the final manuscript. BJ: critically revised the manuscript and approved the final manuscript. All authors read and approved the final manuscript before submission.

## Acknowledgments

The authors acknowledge Madda Walabu University, College of Health Sciences staff for their support during this research work.

Additional Files

Appendices A: PRISMA checklist

Appendices B: Examples of search strategy

Appendices C: The risk-of-bias assessment results for included studies

Appendices D: Sensitivity analysis for included studies of BBFs

